# Enhanced real-time mass spectrometry breath analysis for the diagnosis of COVID-19

**DOI:** 10.1101/2023.06.21.23291712

**Authors:** Camille Roquencourt, Hélène Salvator, Emmanuelle Bardin, Elodie Lamy, Eric Farfour, Emmanuel Naline, Philippe Devillier, Stanislas Grassin-Delyle

## Abstract

**Background:** Although rapid screening for and diagnosis of COVID-19 are still urgently needed, most current testing methods are either long, costly, and/or poorly specific. The objective of the present study was to determine whether or not artificial-intelligence-enhanced real-time MS breath analysis is a reliable, safe, rapid means of screening ambulatory patients for COVID-19.

**Methods:** In two prospective, open, interventional studies in a single university hospital, we used real-time, proton transfer reaction time-of-flight mass spectrometry to perform a metabolomic analysis of exhaled breath from adults requiring screening for COVID-19. Artificial intelligence and machine learning techniques were used to build mathematical models based on breath analysis data either alone or combined with patient metadata.

**Results:** We obtained breath samples from 173 participants, of whom 67 had proven COVID-19. After using machine learning algorithms to process breath analysis data and further enhancing the model using patient metadata, our method was able to differentiate between COVID-19-positive and -negative participants with a sensitivity of 98%, a specificity of 74%, a negative predictive value of 98%, a positive predictive value of 72%, and an area under the receiver operating characteristic curve of 0.961. The predictive performance was similar for asymptomatic, weakly symptomatic and symptomatic participants and was not biased by the COVID-19 vaccination status.

**Conclusions:** Real-time, non-invasive, artificial-intelligence-enhanced mass spectrometry breath analysis might be a reliable, safe, rapid, cost-effective, high-throughput method for COVID-19 screening.

## INTRODUCTION

There have been more than 640 million confirmed cases of COVID-19 since the start of the pandemic [1]. The reference diagnostic testing technique is based on the detection of genetic material from severe acute respiratory syndrome coronavirus 2 (SARS-CoV-2) in nasopharyngeal swabs via a reverse transcription polymerase chain reaction (RT-PCR) [2]. This sampling and testing strategy is time-consuming, requires qualified personnel, and involves costly biological consumables. SARS-CoV-2 viruses are shed from the respiratory tract for about 10 days after disease onset in patients with mild COVID-19 and 20∼40 days after disease onset in patients with severe COVID-19 [3]. False-negative test results may occur in up to 20% to 67% of patients, with positive SARS-CoV-2 PCR tests for 93% of bronchoalveolar fluid samples, 72% of sputum samples, 63% of nasal swabs, and 32% of pharyngeal swabs [2, 4]. Rapid antigen detection (RAD) tests for a SARS-CoV-2 infection have also been developed; they are just as specific as RT-PCR assays but are much less sensitive (∼70%) [5]. Hence, the need for non-invasive, reliable, easy-to-use, cost-effective, validated diagnostic and screening tests with a rapid turn-around-time is still major.

Breath analysis is a non-invasive, real-time, point-of-care technique based on the detection of volatile organic compounds (VOCs). The thousands of VOCs in human breath identified to date are related to physiological and pathological processes (e.g. infections and inflammation) [6, 7]. A number of studies have highlighted the value of breath VOC analysis for the diagnosis of COVID-19, using real-time [8–10] and offline [11–13] mass spectrometry (MS), ion mobility spectrometry [14, 15], Fourier-transform infrared spectroscopy [16], surface-enhanced Raman scattering [17], other sensor technologies (electronic noses) [18–20], and detection dogs sniffing sweat samples [21–24]. Artificial intelligence and machine learning techniques have been frequently applied to the field of breath analysis in general and the diagnosis of COVID-19 in particular; support vector machines, principal component analysis, random forests, artificial neural networks, elastic nets and decision trees have been used to set up predictive models for diagnosis or disease classification [8, 13, 18–20]. It is known that MS breath analysis provides high-dimension data. We hypothesized that the additional implementation of clinical metadata in machine learning models might improve the predictive performance. The objective of the present study was therefore to determine whether or not artificial-intelligence-enhanced real-time MS breath analysis is a reliable, safe, rapid means of screening ambulatory patients for COVID-19.

## METHODS

### Study design and participants

We conducted two prospective, open, interventional studies (VOC-COVID-Diag and VOC-SARSCOV-Dep) in a single university hospital (Foch Hospital, Suresnes, France) and sought to assess the value of VOC analysis (using either proton transfer reaction - mass spectrometry (PTR-MS), electronic noses, or detection dogs) in the diagnosis of COVID-19. The two study protocols were registered (VOC-COVID-Diag: EudraCT 2020-A02682-37; VOC-SARSCOV-Dep: EudraCT 2021-A00167-34) and approved by an independent ethics committee. Written, informed consent was obtained from all the participants. The detection dog results have been published elsewhere [21]; here, we report the results of the real-time MS analysis.

The participants were (i) adults (aged 18 or over) who had to be screened for COVID-19 in the emergency department or (ii) healthy adult volunteers (both vaccinated and unvaccinated) free of COVID-19. The main exclusion criterion was pregnancy. The symptoms frequently associated with COVID-19 were used to calculate a COVID-19 symptom score (SS) on a scale of 0 to 4, based on the absence (scored as 0) of nonspecific symptoms (fever, cough, sore throat, malaise, headache, nausea, vomiting, diarrhoea), the presence (scored as 1) of one of these nonspecific symptoms, or the presence of one (scored as 3) or two (scored as 4) of the most typical, predictive COVID-19 symptoms (myalgia, anosmia, ageusia, dyspnoea, or hypoxemia)[21].

### Study measurements and procedures

One breath sample per participant was collected by trained staff wearing surgical gloves and personal protective equipment. Sampling consisted of a single, deep inhalation and then exhalation through a single-use mouthpiece fitted with a non-return valve into a Tedlar^®^ sample bag (SKC Inc., Eighty Four, PA, USA), which had previously been flushed with ultrapure nitrogen. The bag was then hermetically sealed for immediate transport to the analysis room. Measurements were made with a proton-transfer-reaction quadrupole time-of-flight mass spectrometer (Ionicon Analytik GmbH, Innsbruck, Austria), with the following settings: source voltage, 120 V; drift tube pressure, 3·8 mbar; drift tube temperature, 60°C; and drift tube voltage, 959 V. The mass spectrum was acquired up to *m/z* 392, with a time resolution of 1 s. We recorded medical data, including symptoms frequently associated with COVID-19 (fever, cough, dyspnoea, anosmia, ageusia, fatigue, etc.), underlying health conditions, and medications being taken at the time of sampling. Each participant’s COVID-19 status was determined with molecular assays (the Alinity m SARS-CoV-2 RT-PCR assay or ID Now^®^ COVID-19 assay (Abbott, Issy les Moulineaux, France)) of nasopharyngeal swabs. The threshold cycle for a positive RT-PCR had to be below 40. Any history of a SARS-CoV-2 infection in the months before the time of sampling was determined by applying serological assays (COVID-19 BSS^®^, Biosynex, Fribourg, Switzerland).

### Data processing and statistical analysis

The MS data were processed with the *ptairMS* package in R [25]. Room air was analysed before sample measurement, and breath was differentiated from background air using acetone (*m/z* 59.049) as a tracer VOC. Mass calibration was performed every minute, using the peaks at *m/z* 21.022, *m/z* 60.053, *m/z* 203.943 and *m/z* 330.850. After an alignment step, we selected features (i) present in at least 60% of the participants in a given group and (ii) with a statistically significant difference in signal intensity between room air and exhaled breath in at least 30% of the samples. Missing data were then imputed from the raw data; isotopes and saturated ions (*m/*z 37.028 and 59.049) were removed, as were outlier samples (defined as those with Z-score >5 for at least 10% of the features). Next, we applied the normalization method for metabolomics data using optimal selection of multiple internal standards (NOMIS) [26] method using *m/z* 21.022 (the primary ion isotope), *m/z* 39.022 (the water cluster isotope) and *m/z* 55.038 (water trimers) as normalization features. Lastly, the data were log-transformed.

A univariate analysis was performed using Wilcoxon’s signed-rank test and *p*-values adjusted to control for the false discovery rate [27]. In a multivariate analysis of the data, we applied a principal component analysis and then a random forest machine learning algorithm with a stratified five-fold cross validation repeated four times in order to minimise overfitting. We also applied feature selection by backward recursive feature elimination [28] to reduce dimension, improve accuracy and select the more relevant feature. This process iteratively ranks features according to their importance and removes the weakest one until the performance no longer improves from one iteration to the next. Given that the prime objective of high-throughput breath screening is to determine which patients should then undergo gold-standard testing, the model’s decision cut-off value was chosen to optimize the sensitivity of patient classification. The predictive performance was assessed in term of the sensitivity, specificity, negative predictive value (NPV), positive predictive value (PPV) and area under the curve. Potential confounding covariates linked to COVID-19 status were investigated by applying Wilcoxon’s test or principal component analysis within each group, as described previously [9].

## RESULTS

### The study population

Breath samples from 173 participants (included between October 21^th^, 2020 and June 30^th^, 2022) were analyzed with PTR-MS. One participant was considered to be an outlier and so was excluded from the analysis. Of the 172 remaining participants, 106 had a negative RT-PCR test and 67 had a positive RT-PCR test. For the majority of patients, the RT-PCR test was performed on the same day as the breath analysis (median [interquartile range] time interval: 0 [0–3] days]). The negative and positive participants differed with regard to certain demographic and clinical characteristics (Table 1).

**Table 1:** Patient characteristics and treatments.

|  | **COVID -**<br>**(n = 106)** | **COVID +**<br>**(n = 67)** | ***p* value** |
| --- | --- | --- | --- |
| Sex (M/F) | 50/56 | 31/36 | 1 |
| Age | 46 ± 18 | 56 ± 14 | <0.001 |
| Patients / volunteers (n) | 97 / 9 | 67 / 0 | - |
| Presence of: (n (%)) |  |  |  |
| - Fever | 15 (14.2%) | 32 (47.8%) | <0.001 |
| - Cough | 12 (11.3%) | 39 (58.2%) | <0.001 |
| - Dyspnoea | 12 (11.3%) | 34 (50.7%) | <0.001 |
| - Anosmia | 3 (2.8%) | 10 (14.9%) | 0.003 |
| - Ageusia | 1 (0.9%) | 7 (10.4%) | 0.004 |
| - Fatigue | 8 (7.5%) | 30 (44.8%) | <0.001 |
| - Aches | 6 (5.7%) | 17 (25.4%) | <0.001 |
| Symptom score: | 0.32 ± 0.66 | 2.2 ± 1.1 | <0.001 |
| Medical history: (n (%)) |  |  |  |
| - High blood pressure | 12 (11.3%) | 19 (28.4%) | 0.008 |
| - Asthma | 3 (2.8%) | 9 (13.4%) | 0.018 |
| - Overweight | 5 (4.7%) | 8 (11.9%) | 0.144 |
| - Diabetes | 5 (4.7%) | 7 (10.4%) | 0.255 |
| - Organ transplant | 1 (0.9%) | 4 (6.0%) | 0.055 |
| Previous COVID-19 infection | 12 (11.3%) | 0 (0%) | - |
| COVID-19 vaccination: | 79 (74.6%) | 13 (19.4%) | <0.001 |
| Previous treatment with corticosteroids | 5 (4.7%) | 33 (49.3%) | <0.001 |

Continuous data are presented as the mean ± standard deviation.

### Breath analysis

#### Multivariate analysis

Processing of the real-time PTR-MS data yielded 71 features that were reproducibly detected in the exhaled breath of the patient cohort (Supplementary Table 1). In a principal component analysis, a plot of the second and third components suggested that the COVID-negative and COVID-positive samples were at least partially segregated (Figure 1, and loadings plot in Supplementary Figure 1). A base model (consisting of a random forest with 12 features selected from the 71 and a decision cut-off of 0.2) classified the patients with a sensitivity of 94%, a specificity of 70%, a negative predictive value of 95% and a positive predictive value of 67%, with five-fold cross validation repeated four times. The following clinical metadata were then included in the model as explanatory variables: the symptom score, prior corticosteroid treatment (as a binary yes/no variable) and vaccination status (also as a binary variable). The symptom score and corticosteroid treatment were selected by the algorithm and led to an improved final model with 18 VOC features; it classified participants with a sensitivity of 98%, a specificity of 74%, a negative predictive value of 98% and a positive predictive value of 72% (again with a decision cut-off of 0.2). The receiver operating characteristic curves and metrics for the models (breath only or breath + clinical data) are shown in Figure 2.

**Figure 1:**
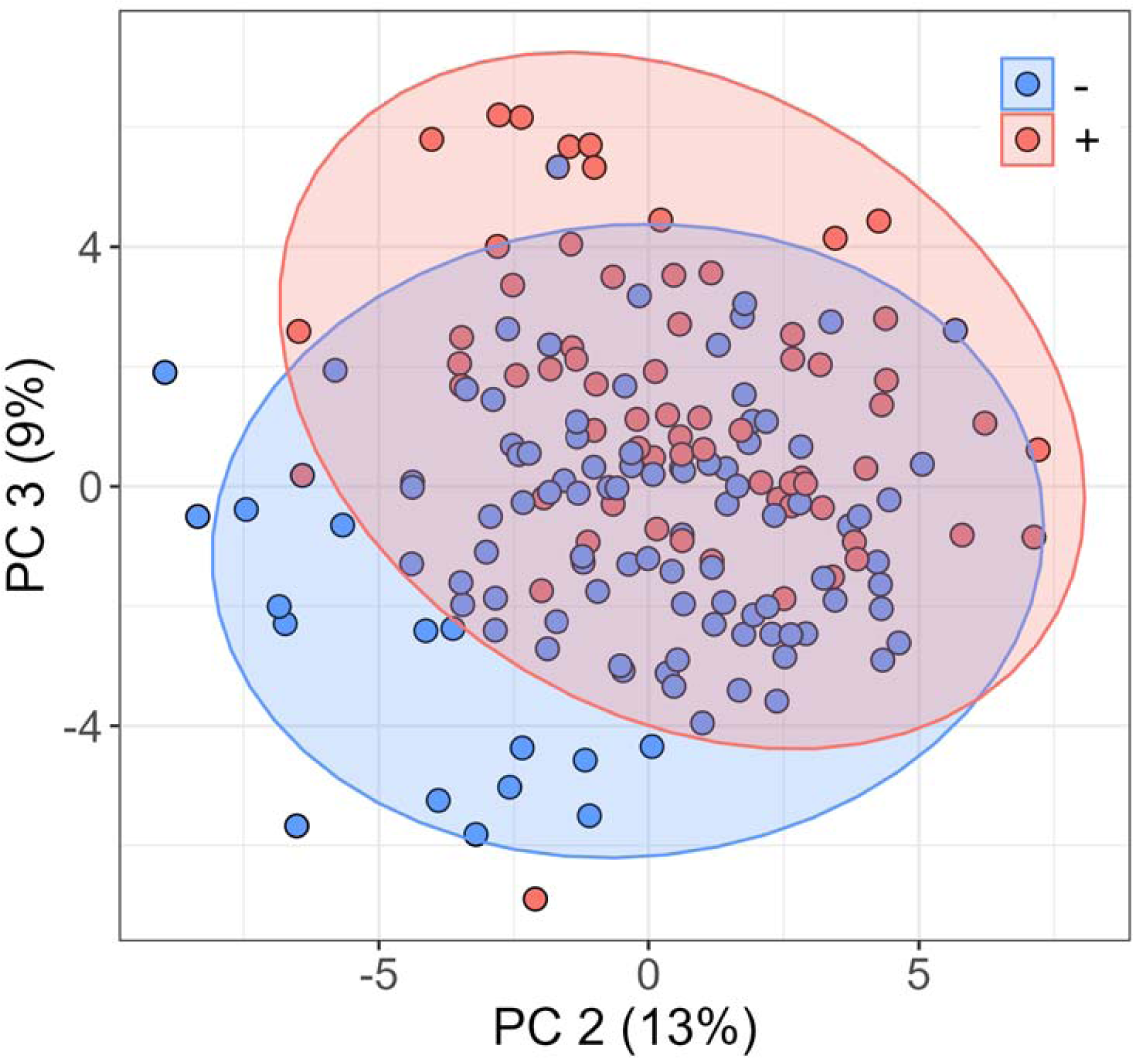
Multivariate analysis. Principal component analysis of the breath signature in participants with a positive (red) or negative (blue) PCR test for SARS-CoV-2.

**Figure 2:**
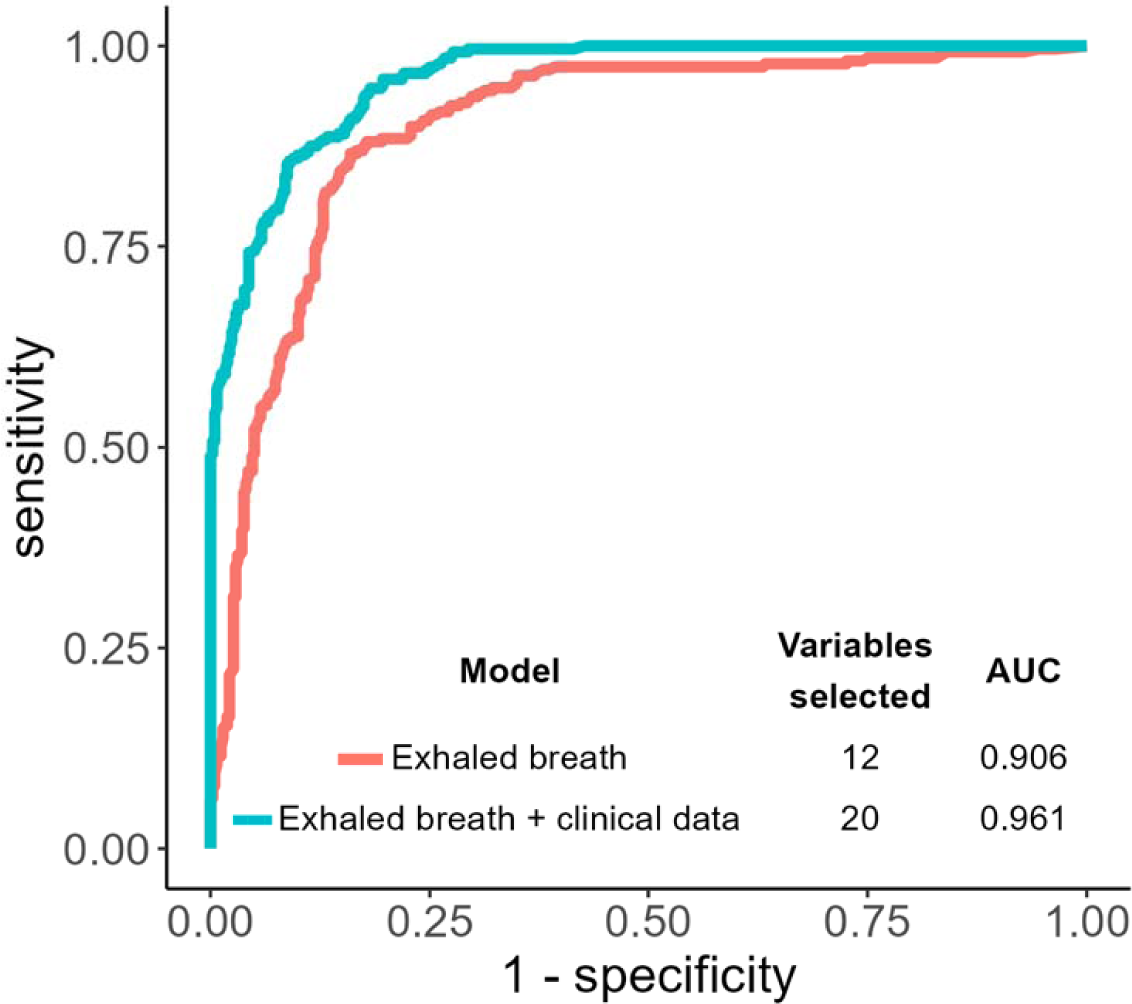
Receiver operating characteristic curve of the basic model (exhaled breath data only) and final model (exhaled breath and medical metadata) with random forest, recursive feature elimination, and NOMIS after five-fold cross validation repeated four times. AUC: area under the curve.

#### Univariate analysis

The univariate analysis highlighted two features that were already part of the multivariate model for differentiating between COVID-19-positive and -negative participants: in COVID-19-positive participants, one feature (*m/z* 99.08) was more intense and the other (*m/z* 63.03) was less intense (Figure 3). We then queried the Human Breathomics Database [29]. Putative annotations for the two compounds are shown in Table 2.

**Figure 3:**
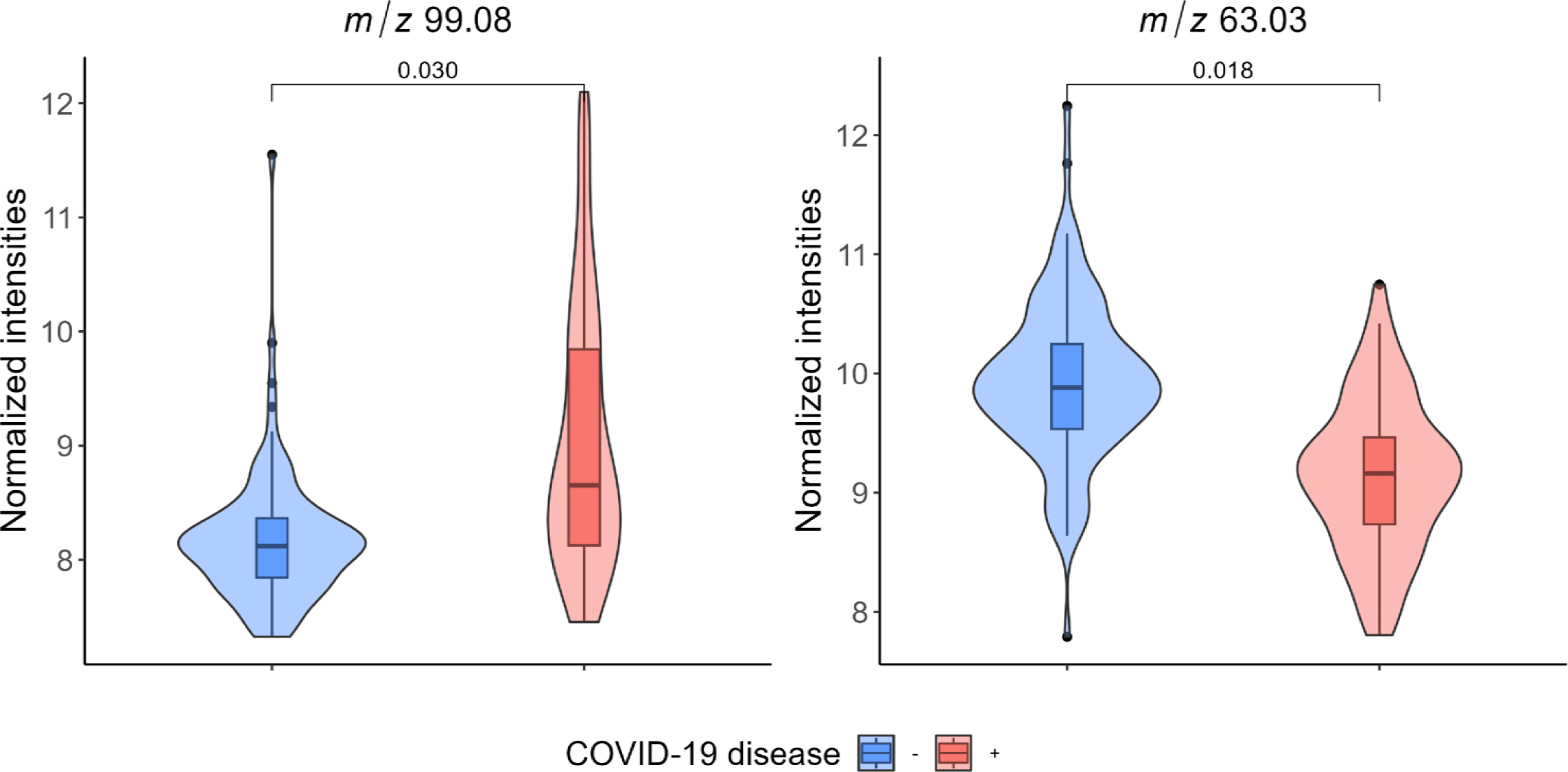
Univariate analysis, showing VOCs whose expression levels differed when comparing COVID-19-negative and -positive groups of participants. The data are expressed as normalised intensities, and *p*-values were calculated with Wilcoxon’s test after correction for the false discovery rate.

**Table 2:** Suggested VOC annotations, after searching the Human Breathomics Database [29].

| VOC (*m/z*) | Suggested molecular formula | Suggested annotation |
| --- | --- | --- |
| 99.08 | $[\text{C}_6\text{H}_{10}\text{O} + \text{H}]^+$ | 2-methylpent-2-enal, 3-methylpent-3-en-2-one, 4-methylpent-2-enal, hex-3-en-2-one, hex-4-en-3-one, 2-methylcyclopentan-1-one, 4-methylpent-3-en-2-one, 4-methylpent-3-enal, cyclohexanone, hex-5-en-2-one |
| 63.03 | $[\text{C}_2\text{H}_6\text{S} + \text{H}]^+$ | Ethanethiol, methylsulfanylmethane |

### Relationship between breath VOCs and disease severity

Firstly, the final model’s predictive performance was similar in asymptomatic or weakly symptomatic patients (symptom score ≤ 1: sensitivity: 98.3%; specificity: 72%) and symptomatic patients (symptom score >1: sensitivity: 97.1%; specificity: 73%). Secondly, in the COVID-19-positive group we explored the relationship between breath concentrations of the two above-mentioned features on one hand and virological and clinical variables on the other. The breath concentrations were not significantly associated with the Ct in the PCR (median (range) Ct = 24 (11–37); r = −0.01 for *m/z* 99.08 and r = 0.2 for *m/z* 63.03). The symptom score was correlated with the breath concentrations of *m/z* 99.08 (r = 0.32, p = 0.009) but not that of *m/z* 63.03 (Figure 4). Lastly, we assessed VOC expression in between ambulatory participants from the present cohort vs. severely ill, intubated, ventilated patients cared for in the ICU from our previous study [9]. The breath VOC concentration of *m/z* 99.08 (but not *m/z* 63.03) was higher in patients with severe disease than in patients with mild disease (Figure 5).

**Figure 4:**
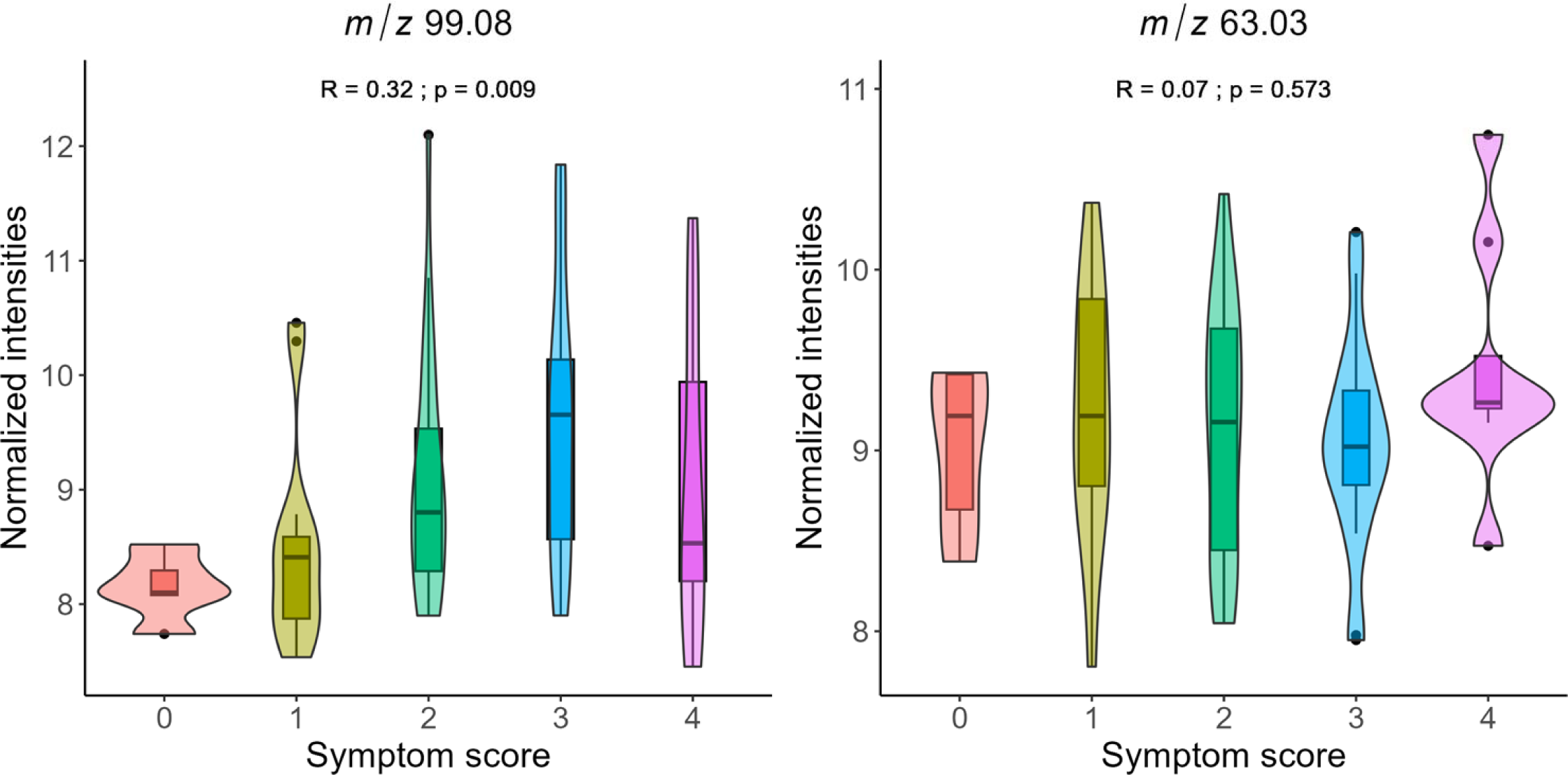
Relationship between the symptom score and the expression levels of *m/z* 99.08 and *m/z* 63.03, as quantified with Spearman’s correlation coefficient. The data are expressed as normalised intensities.

**Figure 5:**
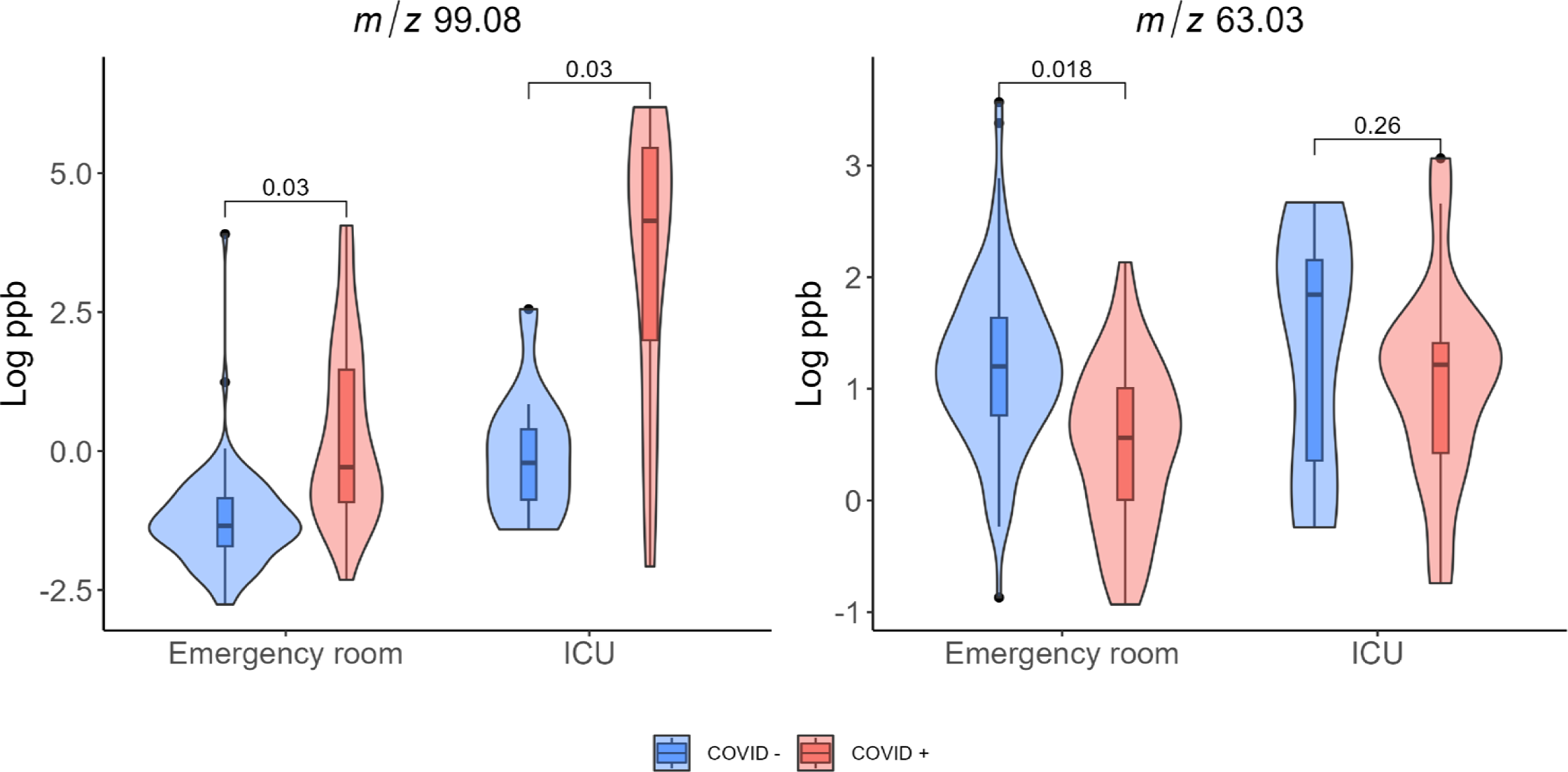
Expression levels of *m/z* 99.08 and *m/z* 63.03 in participants from the emergency department and the ICU. The data are expressed in ppb. The data on patients in the ICU have been published in [9].

### Confounding factors

We sought to rule out potential confounders unrelated to COVID-19, as we had done previously for the oxygen supply in ventilated patients [9]. In the present cohort, prior corticosteroid treatment and COVID-19 vaccination were (as expected) associated with the COVID-19 status and so were investigated. The breath concentration of *m/z* 99.08 was significantly higher in unvaccinated COVID-19-negative participants (n=27) than in vaccinated COVID-19-negative participants (Supplementary Figure 2). In the former group, 6.5 participants (an average after 5-fold cross-validation) would have been incorrectly categorized as COVID-19-positive by the final model and so would have been referred for a reference test for SARS-CoV-2 infection. In all the other groups, there were no confounding effects of COVID-19 vaccination or prior corticosteroid treatment with respect to the concentrations of *m/z* 99.08 and *m/z* 63.03 (Supplementary Figure 2). The absence of a confounding effect was also confirmed for age and sex (Supplementary Figure 3).

## DISCUSSION

Our present results show that real-time MS breath analysis enhanced with clinical metadata and machine learning tools can reliably diagnose COVID-19 in ambulatory subjects. Furthermore, the analysis is non-invasive and has a rapid turn-around time and a low requirement for consumables. Due to their very high sensitivity and negative predictive value, this type of breath analysis might be of value for the rapid, high-throughput, ambulatory identification of COVID-19-negative people; individuals who test positive could be referred for confirmatory testing with a reference method (e.g. a nucleic acid amplification assay). Since these confirmatory tests are invasive and time-consuming, and molecular assay consumables are expensive, the implementation of real-time MS breath analysis would probably have both public health and economic benefits.

With regard to diagnostic performance, the sensitivity of previously reported breath analysis methods for the diagnosis of COVID-19 ranged from 68% to 100% [8-13, 15, 16, 18-20]. Sensitivities greater than 95% were only achieved for methods whose sensor technologies could not identify individual VOCs [16, 17, 19] and/or had analytical runtimes longer than 2 mins (and sometimes even 45 mins) [13, 16, 17, 19]. For some of these techniques, the potential confounding effects of comorbidities, COVID-19 vaccination status and prior therapies have not been studied [16, 17]. When considering the two previous studies of PTR-MS in similar patient populations, one reported the down-regulation of specific VOCs [11] and the other gave a sensitivity of just 81% [8]. These results suggest that the combination of high-resolution MS systems with dedicated software (such as the *ptairMS* package) [25] can markedly improve analytical performance, peak detection, and sample alignment in cohorts of patients. In the above-mentioned studies, the specificity ranged from 75% to 100%. Studies of detection dogs sniffing sweat samples or face masks highlighted excellent diagnostic performances (sensitivities of 61% to 100%, and specificities of 84% to 94%) [21–24], and further strengthened arguments in favour of an odorant signature for COVID-19. However, dog-based testing is limited by the time needed for sweat collection or mask wearing, the need to continuously (re)train the dogs, the presence of between-animal differences and variability over time, and the inability to identify individual VOCs.

Rapid antigen detection tests can also be used to screen for SARS-CoV-2 infection, although their sensitivity with nasopharyngeal swabs ranges from 12% to 98% [4, 5, 30–32]. The lowest sensitivity is observed in asymptomatic or weakly symptomatic patients, which increases the likelihood of false-negative results [4, 5, 30–32]. Since up to 50% of cases can be attributed to transmission from asymptomatic or presymptomatic patients, the high probability of false-negatives might limit the value of RAD tests for mass screening [33, 34]. The predictive performance of our real-time MS breath testing was independent of the viral load and the intensity of the COVID-19 symptoms. Rapid antigen detection tests have a specificity of close to 100% and give few false-positive results; hence, RAD tests are best used to confirm a diagnosis or to differentiate between highly contagious individuals (with a high SARS-CoV-2 load) and less contagious individuals. High-throughput COVID-19 screening in healthcare establishment or busy public trafficked places could perhaps be achieved by combining rapid, sensitive, non-invasive, low-consumable-cost, real-time MS breath testing to quickly identify positive patients for referral to a specific confirmatory RAD test. Importantly, we found that prior corticosteroid treatment and COVID-19 vaccination did not have a significant impact on the diagnosis; in contrast, recent vaccination might interfere with other methods (such as canine olfaction) described in the literature [21].

One important strength of the present breath analysis study was the use of metadata on the patients’ symptoms, vaccination status and medications - all of which can be easily collected at the time of sampling). Hence, the use of artificial intelligence (machine learning) algorithms to simultaneously process breath analysis data and patient metadata improved the mathematical model’s diagnostic performance, relative to models based on breath analysis data alone. Although artificial intelligence tools are now widely used to find diagnostic biomarkers for COVID-19 in breath metabolomic data [8, 13, 18–20], the present study is the first to have included both breath data and easy-to-collect clinical metadata.

In order to ensure that features of interest are detected from exhaled breath and are unrelated to environmental contamination, our study design and data processing workflow included several steps. First, only features with an expression level that differed from room air were considered for statistical analysis. Ambient air for this background removal step was taken from the instrument room, as ambient air from the room where patients stand is contaminated by all VOCs exhaled by the patients. All samples from COVID-19 positive and negative patients were then collected in the same place and processed in the same way, with a similar hospitalisation status/time and in a random order as the analyses were performed at the time the patients arrived at the hospital. This ensures that VOCs from environmental air and any contaminants, which have the same expression level regardless of group, are excluded from the statistical analysis.

With respect to the nature and pathogenic role of VOC biomarkers of COVID-19, the main feature of the present study (*m/z* 99.08) had already been identified in our study of intubated, ventilated patient with COVID-19-related acute respiratory distress syndrome [9]. Xue *et al.* also reported cyclohexanone as a candidate COVID-19 VOC biomarker, which *m/*z for the [M+H]^+^ ion is 99.08 [13]. Our present results showed that the expression level of *m/z* 99.08 was independent of the viral load but not the disease severity. All the suggested annotations for this feature correspond to ketones and aldehydes. These families have already been reported as potential markers of disease states in patients with COVID-19 [10, 11, 13, 15], asthma, or chronic obstructive pulmonary disease [35–38] and after an inhaled endotoxin challenge in healthy volunteers [39]. Although these VOCs and disease states may be interrelated, the underlying biochemistry has not been fully characterized. However, the aldehyde VOC octanal was recently found to be an agonist of the olfactory receptors that are expressed in immune cells (such as macrophages) and are involved in the pathogenesis of the oxidative stress and inflammatory processes in a murine model of atherosclerosis and in human monocyte-derived macrophages [40]. Hence, olfactive receptors might be targeted by certain VOCs in the body and could perhaps be modulated by appropriate pharmacological interventions.

The main limitations of the present study are related to the sample size; our observations will require confirmation in an external validation cohort. Overfitting was however limited during data analysis by both feature selection (71 variables were pre-selected from the 173 patients based on their reproducibility and difference in intensity between exhaled breath and ambient air, and then only 12 where selected by the final machine learning algorithm) and cross-validation. Validation on external cohort would also constitute an opportunity to assess the specificity of the COVID-19 signature, relative to other infectious diseases affecting (or not) the respiratory tract. Although the RT-PCR assay is considered to be the gold standard diagnostic technique and was used here as the comparator, it also has limitations: inappropriate sampling may give false-negative results, whereas a positive result objectifies the presence of viral genetic material in the airways but not necessarily intact (virulent) viruses. Lastly, on the basis of our present data, we were unable to determine the nature or structure of the VOCs of interest; their formal annotation will require the comprehensive use of additional analytical methods.

The present study also had a number of strengths, including the use of high-sensitivity, real-time mass spectrometry capable of quantifying specific VOCs, the implementation of clinical metadata to improve the artificial intelligence algorithm, and the good overall diagnostic performance (especially the excellent negative predictive value). In contrast to classical GC-MS instruments, PTR-MS is easy to implement in a clinical setting as it only requires a power supply and water. Breath analysis for COVID-19 diagnosis with PTR-MS analysis may be cost-effective for several reasons, although COVID-19 screening techniques and strategies (and therefore the associated costs) are highly dependent on the organisational structure (screening centres, outpatient clinics, other healthcare establishments, etc.). For example, COVID-19 RT-PCR tests may be carried out on almost fully automated nucleic acid extraction and amplification systems, with up to 250 runs per day. Such instrument costs around two hundred thousand euros, plus around €20 per sample for consumables and personnel. On the other hand, PTR-MS technology is not widely used; in the absence of economies of scale, the cost of an high-resolution, high-sensitivity instrument may be twice as much, but the per-test cost of consumables is just a few euro cents and the analysis of a patient’s sample takes 1 minute, making it possible to analyse several tens patients a day. Hence, if we take the analysis of 10,000 samples as an example, the analysis cost would be approximately the same for RT-PCR and PTR-MS, but analysis by PTR-MS would take around 2 times less time than with RT-PCR.

Taken as a whole, our results established a specific breath metabolomic signature which, when combined with clinical metadata, allowed reliable, non-invasive, high-throughput COVID-19. We now intend embed all the hardware control and artificial intelligence tools for breath and online data analysis in a user-friendly, automated software package so that staff with basic training can screen for COVID-19 in less than 1 minute per person. This set-up could be used in subsequent validation and extension studies. A non-invasive breath analysis workflow with low consumable use (disposable mouthpieces only) and a rapid turn-around time might have health economic advantages over existing methods by rapidly identifying cases, halting the spread of the virus, and enabling the provision of appropriate care to ill people.

## Data Availability

The study protocol and the datasets generated during and/or analysed during the current study, including deidentified participant data, will be available (after publication) from the corresponding author on reasonable request.

## ACKNOWLEDGEMENTS

The authors thank all the staff members in the emergency department and the clinical research team involved in the study. We also thank Dr David Fraser (Biotech Communication SARL, Ploudalmézeau, France) for copy-editing assistance.

## CONTRIBUTORS

S.G.D. and P.D. conceived the study. P.D., E.F. and S.G.D. collected epidemiological, laboratory and clinical data. H.S. assisted in patient recruitment and clinical research. E.B., E.L. and E.N. contributed to analytical experiments and data acquisition. C.R. and S.G.D. developed software and analysed the data. S.G.D. and C.R. drafted the manuscript. H.S., E.B., E.L., E.F., E.N., P.D. revised the manuscript. All authors read and approved the final version of the manuscript.

