## Supplementary Material for "Enhanced real-time mass spectrometry breath analysis for the diagnosis of COVID-19"

**SUPPLEMENTARY TABLE**

**Supplementary Table 1: Features detected with PTR-MS breath analysis.**

| ***m/z*** | **COVID -**  **(*n* = 106)** | **COVID +**  **(*n* = 67)** |
| --- | --- | --- |
| 99.08 | 0.40 (0.29) | 0.76 (1.85) |
| 63.03 | 3.09 (2.68) | 1.89 (1.81) |
| 43.02 | 38.1 (26.2) | 43.4 (42.5) |
| 77.06 | 3.54 (3.15) | 2.64 (3.41) |
| 63.01 | 1.85 (1.69) | 1.20 (1.54) |
| 31.02 | 105.2 (89.9) | 89.6 (130.4) |
| 137.18 | 0.05 (0.07) | 0.04 (0.07) |
| 58.04 | 0.56 (0.43) | 0.56 (0.63) |
| 31 | 3.92 (1.33) | 3.69 (1.69) |
| 84.94 | 0.02 (0.04) | 0.03 (0.04) |
| 287.24 | 0.02 (0.03) | 0.03 (0.02) |
| 107.05 | 0.29 (0.14) | 0.33 (0.13) |
| 51.04 | 1.17 (1.08) | 1.01 (0.77) |
| 117.09 | 0.56 (0.54) | 0.72 (0.67) |
| 27.99 | 3.19 (0.5) | 3.13 (0.47) |
| 67.05 | 1.73 (1.06) | 2.23 (1.14) |
| 137.06 | 0.07 (0.06) | 0.08 (0.05) |
| 33.03 | 200.7 (160.4) | 179.6 (125.4) |
| 27.99 | 3.18 (0.46) | 3.12 (0.57) |
| 34.04 | 2.11 (1.95) | 2.09 (1.33) |
| 38.01 | 0.54 (0.46) | 0.71 (0.46) |
| 41.04 | 162.9 (102.5) | 197.1 (126.1) |
| 42.01 | 2.72 (1.3) | 3.04 (1.89) |
| 42.04 | 4.79 (7.37) | 7.18 (6.26) |
| 43.99 | 16.2 (5.2) | 15.0 (5.5) |
| 44.02 | 0.53 (0.59) | 0.8 (1.14) |
| 45.00 | 764.2 (134.4) | 737.9 (170.3) |
| 45.03 | 61.0 (50.6) | 83.3 (58.8) |
| 46.04 | 0.79 (0.89) | 1.21 (1.32) |
| 49.01 | 0.67 (1.6) | 0.55 (1.48) |
| 49.03 | 0.10 (0.11) | 0.11 (0.14) |
| 53.00 | 0.46 (0.28) | 0.56 (0.47) |
| 57.03 | 1.54 (0.98) | 1.81 (1.16) |
| 59.05 | 1461.9 (1365.5) | 1222.9 (1627.1) |
| 59.07 | 117.7 (117.6) | 100.7 (183.6) |
| 60.05 | 41.5 (36.2) | 37.5 (51.2) |
| 61.05 | 3.35 (2.96) | 3.22 (3.84) |
| 62.03 | 0.83 (0.35) | 0.76 (0.43) |
| 68.06 | 0.54 (0.31) | 0.7 (0.46) |
| 69.07 | 64.2 (37.2) | 75.0 (39.0) |
| 69.09 | 5.83 (3.34) | 7.48 (4.45) |
| 70.07 | 3.16 (2.13) | 3.99 (2.58) |
| 78.06 | 0.13 (0.1) | 0.09 (0.13) |
| 79.05 | 0.58 (0.24) | 0.6 (0.26) |
| 81.04 | 0.14 (0.13) | 0.11 (0.06) |
| 81.07 | 2.11 (2.81) | 2.58 (2.91) |
| 82.07 | 0.16 (0.22) | 0.19 (0.19) |
| 87.04 | 2.31 (1.9) | 2.55 (2.69) |
| 87.08 | 0.80 (0.65) | 0.98 (0.65) |
| 89.06 | 1.29 (1.22) | 2.42 (3.51) |
| 91.06 | 0.74 (0.54) | 0.70 (0.35) |
| 92.06 | 0.09 (0.05) | 0.08 (0.06) |
| 93.07 | 0.99 (1.18) | 0.90 (0.91) |
| 100.04 | 0.02 (0.01) | 0.02 (0.02) |
| 100.08 | 0.09 (0.05) | 0.15 (0.16) |
| 105.09 | 0.04 (0.07) | 0.04 (0.06) |
| 107.09 | 1.44 (0.89) | 1.52 (0.71) |
| 108.09 | 0.14 (0.08) | 0.16 (0.07) |
| 110.97 | 0.08 (0.05) | 0.07 (0.04) |
| 113.03 | 0.06 (0.05) | 0.06 (0.05) |
| 115.11 | 0.11 (0.07) | 0.11 (0.06) |
| 118.09 | 0.04 (0.04) | 0.05 (0.05) |
| 137.13 | 0.89 (1.19) | 0.92 (1.22) |
| 142.07 | 0.09 (0.04) | 0.11 (0.04) |
| 223.07 | 0.07 (0.03) | 0.07 (0.04) |
| 225.05 | 0.05 (0.02) | 0.05 (0.02) |
| 297.08 | 0.04 (0.02) | 0.04 (0.02) |
| 299.06 | 0.06 (0.02) | 0.06 (0.02) |
| 300.06 | 0.02 (0.01) | 0.02 (0.01) |
| 15.02 | NQ | NQ |
| 18.01 | NQ | NQ |

Data are presented as median (interquartile range). The top 18 lines (in blue) correspond to the features retained in the final model. NQ: not quantified in ppb (*m/z* < 21.022).

**SUPPLEMENTARY FIGURES**

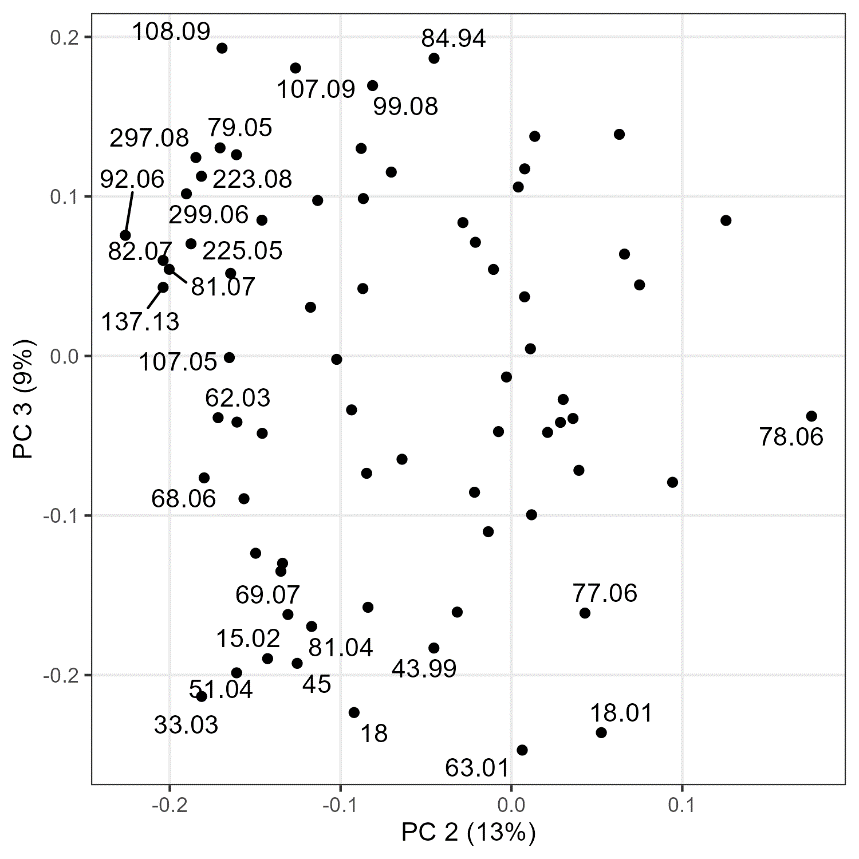

**Supplementary Figure 1:** Loadings plot of the principal component analysis (PCA) model.

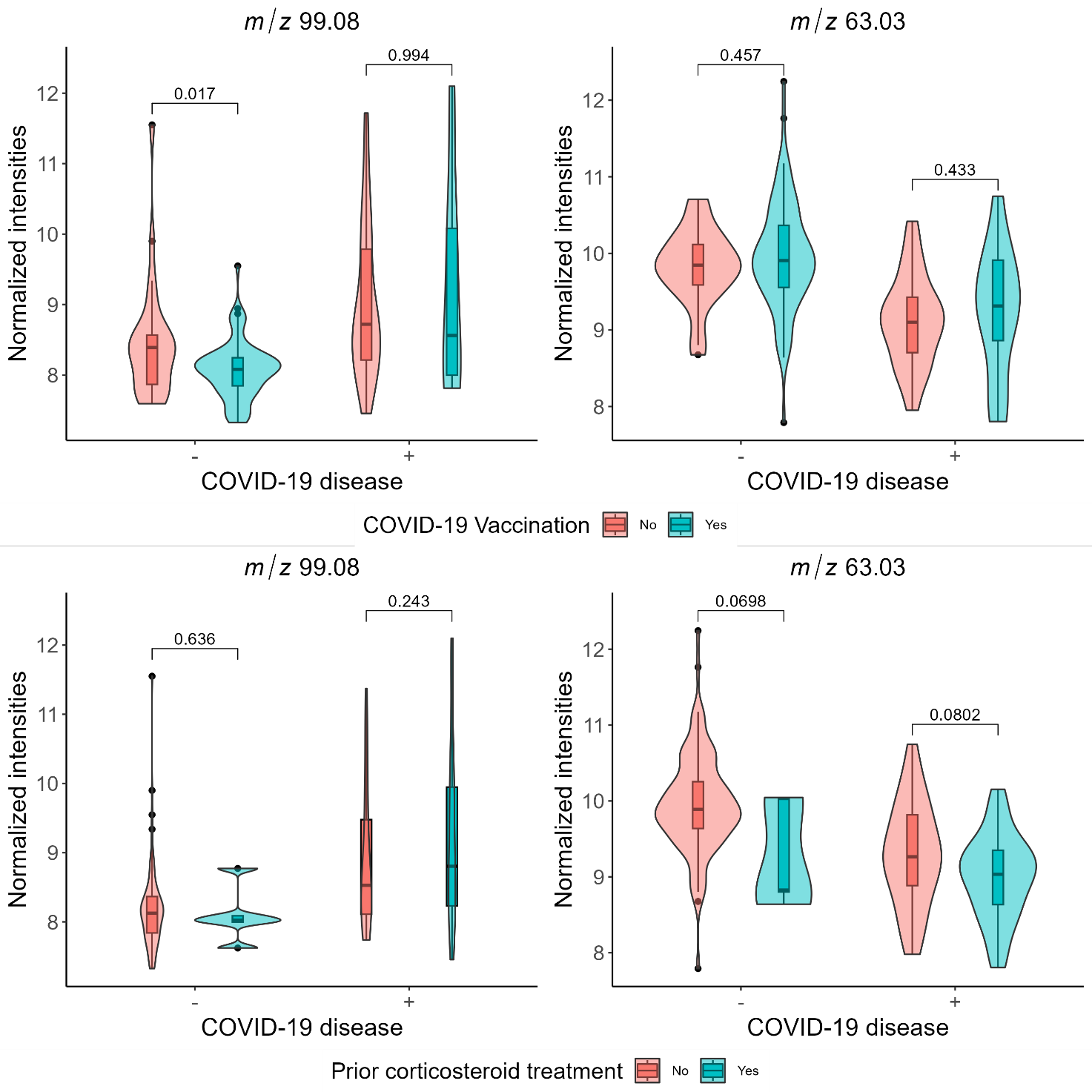

**Supplementary Figure 2:** Effect of confounding factors (COVID-19 vaccination and prior corticosteroid treatment) on the expression levels of *m/z* 99.08 and *m/z* 63.03. Data are expressed as normalised intensities, and *p*-values were calculated with Wilcoxon’s test after correction for the false discovery rate.

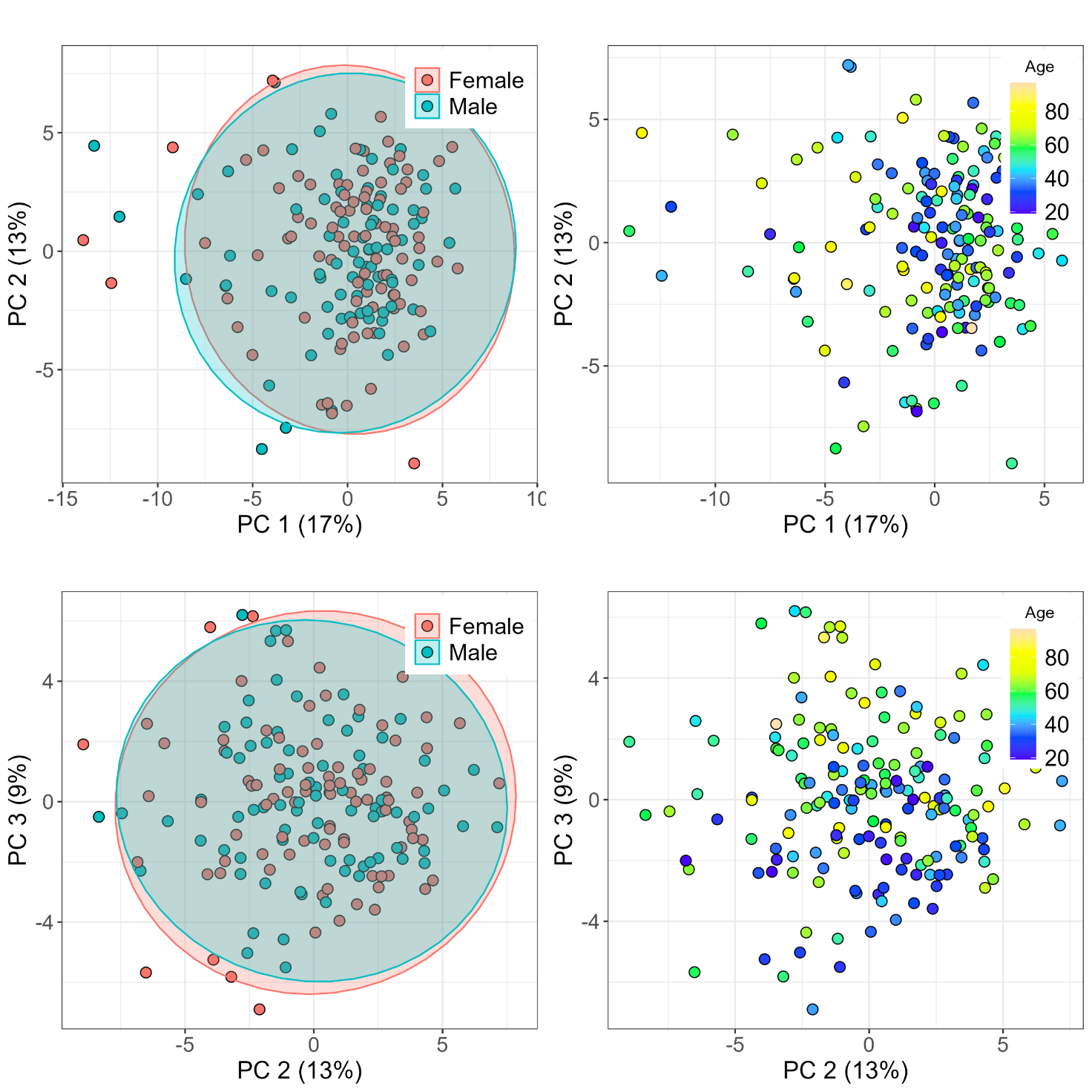

**Supplementary Figure 3:** Effect of confounding factors (age and sex). Principal component analysis of the breath signature in participants by sex (left panels) and age (right panels) for the first and second components (upper panels) and the second and third components (lower panels).
